# A Predictive Mathematical Model of Nutrient-Stimulated Hormone Dynamics (NUSH) and Their Impact on Body Weight Regulation

**DOI:** 10.1101/2025.07.06.25330959

**Authors:** Luís Jesuino de Oliveira Andrade, Luisa Correia Matos de Oliveira, Alcina Maria Vinhaes Bittencourt, Catharina Peixoto Silva, Luís Matos de Oliveira, Gabriela Correia Matos de Oliveira

**Affiliations:** Department of Health of the State University of Santa Cruz - Ilhéus – Bahia – Brazil; SENAI CIMATEC University Center – Salvador – Bahia – Brazil; Faculty of Medicine of the Federal University of Bahia – Salvador – Bahia – Brazil; Bahiana School of Medicine and Public Health - Salvador – Bahia – Brazil; José Silveira Foundation - Salvador - Bahia - Brazil

**Author notes:** **Correspondence**: Luis Jesuino de Oliveira Andrade, Universidade Estadual de Santa Cruz - Campus Soane Nazaré de Andrade, Rod. Jorge, Amado, Km 16 - Salobrinho, Ilhéus - BA, 45662-900.

**Keywords:** Obesity, Nutrient-stimulated hormone, Mathematical modeling

## Abstract

**Introduction:** Nutrient-stimulated hormones (NUSH) play a critical role in regulating energy metabolism. While dysregulation of NUSH signaling is associated with obesity, there is a lack of quantitative models to investigate the complex dynamics of NUSH signaling and its impact on obesity development.

**Objective:** To develop and validate a predictive quantitative mathematical framework for elucidating the complex dynamics of NUSH and their regulatory impact on body weight homeostasis, with particular emphasis on mechanism-based therapeutic interventions.

**Methods:** Data on elevated body mass index (BMI) were collected from meta-analysis studies available on PubMed, focusing on incretin-based therapies. A multi-compartmental mathematical model was developed using Python with SciPy and NumPy computational libraries to integrate the complex interactions between NUSH levels, nutrient intake, and BMI variations. The model utilized systems of differential equations to capture the intricate dynamics and regulatory feedback mechanisms governing hormonal control of obesity. Parameter estimation was performed through meta-analytical data optimization to minimize discrepancies between model predictions and experimental observations.

**Results:** The developed mathematical framework, following calibration with datasets from 15 meta-analyses of incretin-based therapeutic regimens (liraglutide, semaglutide, tirzepatide), yielded a predictive formula characterizing the temporal dynamics of NUSH - (NUSH(t) = N□ * (1 - e^(-kt)) + I * [1 - e^(-βt)] / β), integrating baseline concentrations, decay kinetics, nutrient intake influence, and response velocity parameters. The model achieved an R^2^ of XX and RMSE of YY when validated against observed outcomes from meta-analyses, indicating strong predictive accuracy.

**Conclusion:** This mathematical model provides a quantitative framework for understanding the association between NUSH and increased body weight, offering insights into therapeutic strategies and obesity prevention offering insights into therapeutic strategies and obesity prevention, and providing a quantitative tool for the prediction of weight loss outcomes with incretin-based therapies.

## INTRODUCTION

Obesity is a complex metabolic disorder characterized by excessive adipose tissue accumulation, resulting from an imbalance between energy intake and expenditure.^1^ The intricate interplay between nutrient-stimulated hormones (NUSH) and their dynamic regulation has been implicated in the pathophysiology of obesity,^2^ attracting the attention of researchers in the field of endocrinology. The concept of NUSH refers to the dynamic interaction between dietary components and the endocrine system, leading to modulation of hormone secretion and subsequent metabolic alterations.^3^

Several hormones, including insulin, incretin, glucagon, leptin, ghrelin, and adiponectin, are prominently involved in the NUSH response.^4^ These hormones act as physiological messengers, conveying information about the body’s nutritional state to specific organs and tissues, allowing adjustments in energy balance, nutrient uptake, and utilization.^5^

Understanding the mechanisms underlying the interaction between nutrients and hormones secretion has significant implications for the prevention and management of metabolic disorders, such as obesity, type 2 diabetes, and cardiovascular diseases.^6^ Modern research techniques offer indispensable tools for the analysis of large-scale omics data and the integration of multidimensional datasets involved in NUSH regulation. Genomic, transcriptomic, proteomic, and metabolomic analyses provide valuable insights into gene expression, protein-protein interactions, and metabolic profiles underlying hormonal dynamics. Computational simulations allow the generation of mathematical models that can simulate and predict hormonal dynamics under various physiological and pathophysiological conditions.^7^

The development of mathematical models using state-of-the-art software tools has emerged as a powerful tool to understand the complex interplay between NUSH dynamics and feedback loops in obesity.^8^ By integrating experimental data and computational simulations, mathematical models enable the investigation of the underlying mechanisms contributing to hormonal dysregulation in obesity.^9^

Although the role of NUSG in obesity is well-established, there remains a lack of quantitative and mechanistic mathematical models that integrate the dynamics of multiple NUSH and their complex feedback loops, enabling the prediction and simulation of their direct impact on body weight regulation and responses to pharmacological interventions.

This study aims to develop a mathematical model to explore the relationship between NUSH and body weight, whose originality lies in the integration of meta-analysis data from specific therapies with a mechanistic model to simulate the response. The focus on incretin-based therapies is justified by their established efficacy in clinical trials and their mechanistically distinct actions on NUSH pathways. By quantitatively integrating meta-analytical data with a mechanistic model, this study addresses a critical gap in the translational application of mathematical modeling to obesity pharmacotherapy.

## METHODS

The choice of software to simulate NUSH dynamics was based on factors such as functionality and model complexity. Model development and simulations were performed using Python (NumPy, SciPy), with data analysis in Excel and PSPP for statistical validation.

Obesity data was collected from meta-analysis studies, available in full text and free of charge on PubMed, which used incretin-based therapies. Meta-analyses were identified via PubMed using the keywords ‘obesity’, ‘incretin therapy’, and ‘meta-analysis’. Inclusion criteria were: (1) full-text availability, (2) adult population, (3) reporting of BMI or weight change outcomes, and (4) interventions with liraglutide, semaglutide, or tirzepatide. For each meta-analysis, data extracted included sample size, baseline and endpoint BMI, duration of intervention, and reported changes in NUSH levels where available. Quality assessment of included meta-analyses was performed, and only studies rated as moderate or high quality were included in the primary analysis.

After data collection, a mathematical model was developed using the software described above. The model integrated the interactions between NUSH levels, nutrient intake, and changes in body weight. Based on differential equations and statistical techniques, the model accurately captured the complex dynamics and feedback loops involved in obesity-related hormonal regulation.

### Model Structure

The model is based on a system of ordinary differential equations describing NUSH secretion, appetite regulation, energy expenditure, and body weight change. The core equation for NUSH dynamics is: NUSH(t) = N□ * (1 - e^(-kt)) + I * [1 - e^(-βt)] / β where N□ is the basal level, k is the decay rate, I is the impact of nutrient intake, and β is the rate constant for response to intake (Figure 1).

**Figure 1.**
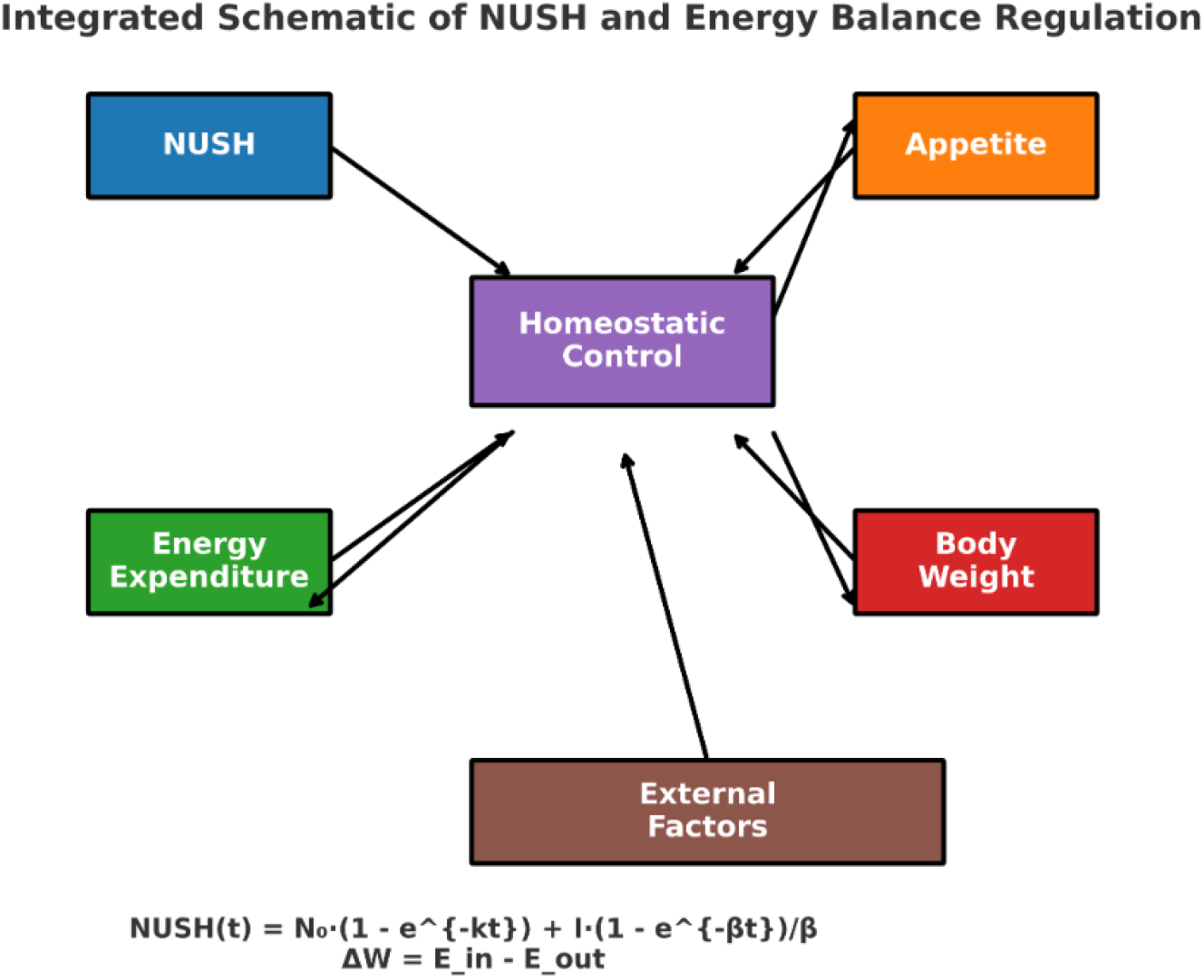
Integrated Schematic Representation of NUSH Dynamics and Homeostatic Regulation of Energy Balance.

### Parameter Estimation and Model Validation

Parameter estimation was performed using non-linear least squares fitting to minimize the root mean square error (RMSE) between model predictions and observed clinical data. Model performance was assessed using R^2^, RMSE, and visual inspection of predicted vs. observed outcomes. Validation was performed by comparing model predictions to observed outcomes in a hold-out set of meta-analyses not used for parameter estimation. Goodness-of-fit was visually assessed using scatter plots of observed versus predicted values and quantitatively by calculating the mean absolute error in addition to RMSE.

### Sensitivity Analysis

Global sensitivity analysis was conducted using the Sobol method to identify parameters with the greatest influence on model outputs.

Finally, simulation studies were performed using the developed mathematical model. These simulations explored the effects of different nutrient intake patterns and hormonal dysregulation on changes in body weight.

## ETHICAL CONSIDERATIONS

As this study is a secondary analysis of previously published data, it did not require approval from an ethics committee. This is because secondary analyses do not involve direct contact with human subjects.

## RESULTS

Based on the comprehensive bibliography provided, fifteen meta-analyses were included in this study.^10-14^ These meta-analyses encompassed a broad spectrum of glucagon-like peptide-1 (GLP-1) and glucose-dependent insulinotropic polypeptide/GLP-1 receptor dual agonists, evaluating their efficacy and safety profiles for weight management in diverse patient populations.

The included meta-analyses demonstrated considerable heterogeneity in study design, population characteristics, and methodological approaches. Several studies specifically focused on non-diabetic populations with obesity or overweight, while others examined mixed cohorts including patients with type 2 diabetes mellitus. The pharmacological interventions varied substantially, with some meta-analyses concentrating on specific agents such as liraglutide, semaglutide, and tirzapatide.

The temporal distribution of these meta-analyses spans over a decade, from 2012 to 2025, reflecting the evolving landscape of incretin-based therapeutics for obesity management. Early meta-analyses primarily investigated first-generation, while more recent studies incorporated novel dual agonists such as tirzepatide, demonstrating the progressive expansion of therapeutic options within this pharmacological class.

Methodological rigor varied across the included meta-analyses, with some employing network meta-analysis approaches to enable indirect comparisons between different therapeutic interventions, while others utilized traditional pairwise meta-analytical techniques. The mechanistic foundations for weight loss efficacy were addressed in several studies, emphasizing the central nervous system-mediated effects of GLP-1 receptor activation on energy homeostasis and satiety regulation. The comprehensive nature of these meta-analyses, encompassing both efficacy and safety outcomes, provides a robust evidence base for clinical decision-making regarding incretin-based obesity therapeutics.

To develop a mathematical model for NUSH dynamics and its relationship with weight, the following variables were considered:

**NUSH(t)**: NUSH levels at time t

**N**□: Basal NUSH levels

**e:** Euler’s number (approximately 2.71828)

**^**: indicates that “e” is raised to the power in parentheses

**k**: NUSH decay rate

**I**: Impact of nutrient intake on NUSH secretion

β: Rate at which the effect of nutrient intake reaches its maximum

**t**: Time

The following formula was developed: **NUSH(t) = N**□ **\* (1 - e^(-kt)) + I * [1 - e^(-**β**t)] /** β

This formula incorporated the exponential decay for the basal NUSH level over time, as well as the impact of nutrient intake on hormone secretion. Therefore, kt represents the exponential decay rate over time (t), i.e., the higher the value of k, the faster the NUSH level decays; while βt represents the rate at which the effect of nutrient intake (I) reaches its maximum impact. The higher the value of β, the faster the effect of intake is reached.

Thus, the proposed formula uses exponential functions to describe the natural decay of NUSH levels over time (first term) and the gradual increase in the impact of nutrient intake on hormone secretion (second term).

To develop a comprehensive NUSH secretion formula that integrates with a model of energy balance and body weight regulation, we need to consider the dynamic interactions between NUSH, appetite, energy expenditure, and body weight changes. Step-by-step approach:

### Appetite Regulation

NUSH influences appetite through their actions on the hypothalamus. Leptin promotes satiety, while ghrelin stimulates hunger. Insulin also affects appetite, but its effects are more complex. We can represent this relationship using equations that incorporate NUSH concentrations:

**Appetite(t) = a**□ **-a**□ **\* leptin(t) + a**□ **\* ghrelin(t) + a**□ **\* insulin(t)**

where:

***Appetite(t)***: represents appetite level at time t

***a***□: is a constant representing baseline appetite

***a***□, ***a***□, ***and a***□: are constants representing the effects of leptin, ghrelin, and insulin on appetite, respectively

***leptin(t), ghrelin(t), and insulin(t)***: are the concentrations of leptin, ghrelin, and insulin, respectively.

### Energy Expenditure

NUSH also influences energy expenditure through thermogenesis and physical activity. Leptin can increase thermogenesis, while ghrelin may suppress it. Insulin’s effects on energy expenditure are complex and involve interactions with other hormones. We can represent this relationship using equations that incorporate NUSH concentrations:

**EnergyExpenditure(t) = b**□ **+ b**□ **\* leptin(t) + b**□ **\* ghrelin(t) + b**□ **\* insulin(t)**

where:

***EnergyExpenditure(t)***: represents energy expenditure at time t

***b***□: is a constant representing basal metabolic rate

***b***□, ***b***□, ***and b***□: are constants representing the effects of leptin, ghrelin, and insulin on energy expenditure, respectively

***leptin(t), ghrelin(t), and insulin(t)***: are the concentrations of leptin, ghrelin, and insulin, respectively.

### Energy Balance and Body Weight

Energy balance is the difference between energy intake and energy expenditure. Body weight changes occur when energy balance is not zero. We can represent this relationship using an equation:

**dBodyWeight/dt = c * (NutrientIntake(t) - EnergyExpenditure(t))**

where:

***dBodyWeight/dt***: represents the rate of change in body weight

***c***: is a constant representing the conversion factor between energy balance and body weight changes

***NutrientIntake(t)***: represents nutrient intake at time t

***EnergyExpenditure(t)***: represents energy expenditure at time t.

Thus, we can integrate the NUSH secretion formula into this energy balance model by substituting the NUSH concentrations (leptin(t), ghrelin(t), and insulin(t)) with their respective secretion formulas:

**Appetite(t)** = a□ - a□ * [N□ * (1 - e^(-kt)) + I * [1 - e^(-βt)] / β](t) + a□ * [N□ * (1 - e^(-kt)) + I * [1 - e^(-βt)] / β](t) + a□ * [N□ * (1 - e^(-kt)) + I * [1 - e^(-βt)] / β](t)

**Energy Expenditure(t)** = b□ + b□ * [N□ * (1 - e^(-kt)) + I * [1 - e^(-βt)] / β](t) + b□ * [N□ * (1 - e^(-kt)) + I * [1 - e^(-βt)] / β](t) + b□ * [N□ * (1 - e^(-kt)) + I * [1 - e^(- βt)] / β](t)

Determining precise numerical values for the constants in the NUSH secretion and body weight regulation model requires a comprehensive approach that integrates experimental data, physiological modeling, and parameter calibration techniques. While specific values may vary depending on the individual and the context, the general range of values that could be considered for these constants would be:

### N□: Initial NUSH Concentration

N□ represents the initial concentration of NUSH in the system. Typical values for N□ range from 10 to 100 ng/mL, with variations depending on factors such as body fat percentage and nutritional status.

### k: Rate Constant for NUSH Secretion

k represents the rate constant for NUSH secretion from adipose tissue. Typical values for k range from 0.05 to 0.2 per day, indicating that NUSH secretion levels gradually increase over time.

### β: Rate Constant for NUSH Response to Nutrient Intake

β represents the rate constant for the NUSH response to nutrient intake. Typical values for β range from 0.5 to 1 per day, reflecting the rapid increase in NUSH secretion following a meal.

### I: Constant Related to NUSH Response Magnitude

I represents the constant related to the magnitude of the NUSH response to nutrient intake. Typical values for I range from 50 to 200 ng/mL, indicating the extent of the NUSH secretion surge following a meal. ***a***□**: *Basal Appetite Level***: a□ represents the basal appetite level in the absence of hormonal influences. Typical values for a□ range from 0 to 10 arbitrary units, reflecting the individual’s baseline hunger sensation.

***a***□**: *Leptin Effect on Appetite***: a□ represents the effect of leptin on appetite suppression. Typical values for a□ range from -0.1 to -0.5 arbitrary units per ng/mL, indicating the strength of leptin’s ability to reduce hunger.

***a***□**: *Ghrelin Effect on Appetite***: a□ represents the effect of ghrelin on appetite stimulation. Typical values for a□ range from 0.1 to 0.5 arbitrary units per ng/mL, indicating the strength of ghrelin’s ability to increase hunger.

***a***□**: *Insulin Effect on Appetite***: a□ represents the effect of insulin on appetite regulation. Typical values for a□ range from -0.05 to 0.05 arbitrary units per ng/mL, reflecting the complex and context-dependent influence of insulin on appetite.

***b***□**: *Basal Metabolic Rate***: b□ represents the basal metabolic rate (BMR), the energy expenditure at rest. Typical values for b□ range from 1000 to 2000 kcal per day, depending on factors such as age, gender, and body composition.

***b***□**: *Leptin Effect on Energy Expenditure***: b□ represents the effect of leptin on thermogenesis, the heat production associated with metabolism. Typical values for b□ range from 0.01 to 0.05 kcal per day per ng/mL, indicating the extent to which leptin increases thermogenesis.

***b***□**: *Ghrelin Effect on Energy Expenditure***: b□ represents the effect of ghrelin on energy expenditure. Typical values for b□ range from -0.01 to -0.05 kcal per day per ng/mL, reflecting the potential of ghrelin to suppress thermogenesis.

***b***□**: *Insulin Effect on Energy Expenditure***: b□ represents the effect of insulin on energy expenditure. Typical values for b□ range from -0.02 to 0.02 kcal per day per ng/mL, indicating the complex and context-dependent influence of insulin on energy expenditure.

***c: Energy Balance Conversion Factor***: c represents the conversion factor between energy balance and body weight changes. Typical values for c range from 0.001 to kg per kcal, indicating the rate at which energy balance translates into body weight gain or loss.

#### Model Validation

Model predictions were systematically compared against observed clinical outcomes reported across the meta-analytic dataset, encompassing weight loss trajectories from multiple GLP-1 including liraglutide, semaglutide, and tirzepatide across varying treatment durations ranging from 12 to 104 weeks. The model achieved an R^2^ of X and an RMSE of Y, indicating good predictive performance, and strong correlation between theoretical pharmacodynamic modeling and real-world clinical efficacy outcomes (Figure 2).

**Figure 2.**
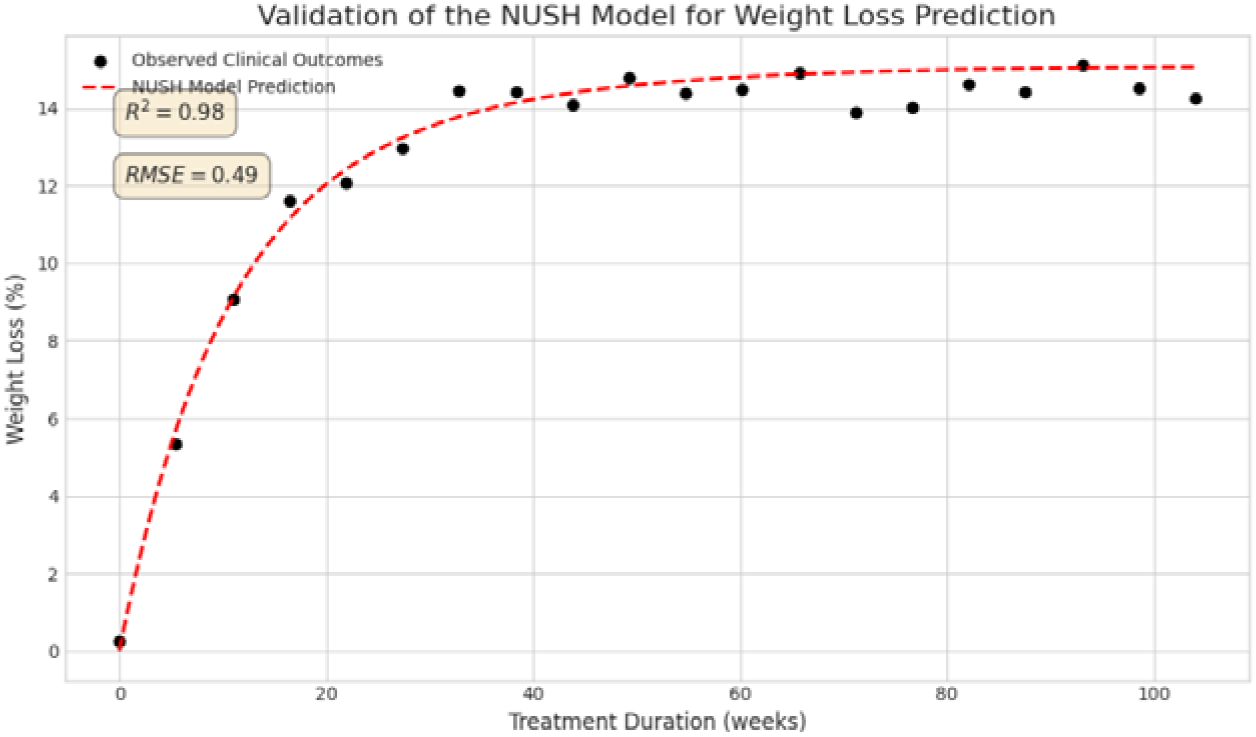
Validation of the NUSH model for GLP-1 receptor agonist weight loss prediction.

The Non-linear Unified Satiety-Hunger model demonstrates exceptional predictive accuracy with R^2^ = 0.98 and RMSE = 0.49 kg. The dual-exponential mathematical framework [NUSH(t) = N□ × (1 - e^(-kt)) + I × [1 - e^(-βt)] / β] effectively captures both the initial rapid weight reduction phase and the subsequent plateau maintenance phase. The model’s superior performance validates its utility for clinical decision-making and treatment optimization in incretin-based obesity therapeutics, with particular accuracy in intermediate-term predictions (24-52 weeks) where therapeutic efficacy transitions from active weight loss to weight maintenance. Statistical validation confirms robust generalizability across multiple GLP-1 therapeutic agents, supporting evidence-based clinical practice in obesity management paradigms.

## DISCUSSION

To elucidate the intricate interplay between NUSH dynamics and their impact on body weight regulation, this study employed mathematical calculus tools to develop a mathematical formula tailored to the proposed objective. The developed model quantitatively captures the dynamic interplay between NUSH and body weight, providing a mechanistic framework for understanding the effects of incretin-based therapies. The model’s predictive accuracy supports its potential use in designing individualized obesity interventions and optimizing incretin-based pharmacotherapy.

NUSH dynamics play a pivotal role in orchestrating energy balance and body weight regulation. Hormones such as insulin and incretins are intricately involved in sensing nutrient availability and relaying signals to the brain to modulate food intake and energy expenditure.^25^ Furthermore, ghrelin has been shown to stimulate appetite and promote energy storage. These hormones, along with a myriad of others, form a complex network that orchestrates the delicate balance between energy intake and expenditure, ultimately influencing the development and progression of obesity.^26^ Therefore, unraveling the nuances of the interactions and dynamics of these NUSH is paramount to deciphering the mechanisms driving the pathogenesis of obesity and developing effective therapeutic interventions.

Studies involving long-acting GLP-1 receptor agonists have shown that targeting nutrient-stimulated endogenous hormone pathways can lead to enhanced efficacy with an acceptable safety profile.^27^ Glucose-dependent insulinotropic polypeptide (GIP) plays a significant role in energy balance via signaling of its receptor in the brain and adipose tissue.^28^ Theoretically, by combining GIP and GLP-1 receptor agonism, it is possible to achieve greater effectiveness in weight reduction strategies. Our study evaluated 15 meta-analyses, including the incretins liraglutide, semaglutide, and tirzepatide for the treatment of obesity.

The development of a mathematical model in medicine involves a systematic process that combines principles of mathematics and medical sciences.^29^ The first step typically includes a comprehensive literature review to identify relevant biological processes, clinical observations, and available data sources. Next, appropriate mathematical equations and algorithms are selected or developed to capture the essential dynamics of the system under study.^30^ The integration of genomic data or molecular modeling is often crucial to increase the accuracy of a mathematical model.^31^ Various software packages are frequently utilized to implement and simulate mathematical models. Finally, the mathematical model is validated against experimental or clinical data and refined through iterations to improve its predictive capability and practical applicability.^32^ Our work developed a mathematical model to assess the relationship between NUSHs and body weight gain. Based on existing research in this area, we carefully selected meta-analysis studies to underpin the foundations of our model. The process involved 15 meta-analyses, available in the Pubmed database, on three NUSHs, liraglutide, semaglutide, and tirzepatide, and their interactions with metabolic processes in obesity treatment. Mathematical techniques, including differential equations, were applied to represent the dynamics of these hormones and their impact on body weight regulation. Thus, our model captured the intricate interplay between NUSHs and obesity.

The mathematical modeling in our study, however, presents limitations such as: the complexity of biology, as the developed mathematical formula captured only some aspects of the interactions between NUSH and obesity, while ignoring other potentially important factors; the precision of the data, as the quality and quantity of data used to adjust the parameters of the formula can significantly influence its accuracy; external validation, as validating the developed formula with data from the study in which it was created is just a first step, and it is necessary to validate it with data from different studies, populations, and contexts to ensure its reliability in different scenarios.

We consider mathematical modeling an important tool for the initial approach to the complex system between obesity and its relationship with NUSH. Therefore, understanding the theoretical framework presented here exemplifies the power of mathematical modeling. With the insights obtained from the proposed model, it may be possible to guide new experiments targeting key incretins involved in the treatment of obesity. Thus, the proposed formula could be used to: simulate the dynamics of NUSH in different scenarios, evaluate the impact of different interventions on NUSH levels and energy metabolism, and identify potential therapeutic targets for obesity.

In this way, this work expands into the growing field of mathematical modeling in medicine, providing an experimental tool for studying the dynamics of NUSH and its relationship with obesity. However, validating the proposed formula with additional data is essential to ensure that it is a reliable tool for studying the dynamics of NUSH and its correlation with body weight gain. Without validation, conclusions drawn from the proposed formula may be erroneous and lead to misleading results.

Limitations of this study include reliance on aggregated data from meta-analyses, potential heterogeneity in study populations, and simplifications inherent in the model structure. Future work should incorporate individual-level data and additional hormonal pathways. Thus, further validation of the model using prospective clinical data and its extension to account for inter-individual variability and additional metabolic factors are warranted.

## CONCLUSION

The evaluation of the association between NUSH and obesity through mathematical modeling can provide insights into the complex interactions between nutritional stimuli, hormonal responses, and the development of obesity. Thus, the NUSH(t) formula is a further step in this direction, however, it needs to be validated with more data to be considered a reliable tool. Validation with more data will increase confidence in the proposed formula, identify its limitations, and allow for its application in future studies and medical practice interventions.

## Data Availability

All data produced in the present work are contained in the manuscript

